# SARS-CoV-2 Airborne Surveillance Using Non-Powered Cold Traps

**DOI:** 10.1101/2021.01.19.21250064

**Authors:** Sven G. Gehrke, Claudia Förderer, Wolfgang Stremmel

**Affiliations:** Medical Center Baden-Baden, Beethovenstr. 2, 76530 Baden-Baden, Germany

**Keywords:** COVID-19, SARS-CoV-2, Aerosols, Quantitative RT-PCR, Spreading Kinetics, Indoor Hotspots, Public Places

## Abstract

**Background:** COVID-19 pandemic is a worldwide challenge requiring efficient containment strategies. High-throughput SARS-CoV-2 testing and legal restrictions are not effective in order to get the current outbreak under control. Emerging SARS-CoV-2 variants with a higher transmissibility require efficient strategies for early detection and surveillance.

**Methods:** SARS-CoV-2 RNA levels were determined by quantitative RT-PCR in aerosols collected by non-powered cold traps. SARS-CoV-2 spreading kinetics and indoor hotspots could be identified in isolation units and at public places within a high-endemic area. These included an outpatient endoscopy facility, a concert hall, and a shopping mall.

**Results:** Indoor COVID-19 hotspots were found in non-ventilated areas and in zones that are predisposed to a buoyancy (chimney) effect. SARS-CoV-2 RNA in those aerosols reached concentrations of 10^5^ copies/mL. Extensive outdoor air ventilation reliably eliminates SARS-CoV-2 aerosol contamination.

**Conclusions:** The method presented herein could predict SARS-CoV-2 indoor hotspots and may help to characterize SARS-CoV-2 spreading kinetics. Moreover, it can be used for the surveillance of emerging SARS-CoV-2 variants. Due to low costs and easy handling, the procedure might enable efficient algorithms for COVID-19 prevention and screening.

## Introduction

COVID-19 is spreading around the world for more than 12 months. Since Spanish flu 100 years ago, no pandemic lead to a comparable medical and economic disaster. Only 2 weeks after the first report, a new coronavirus SARS-CoV-2 had been identified being responsible for the outbreak of first COVID-19 cases suffering from severe atypical pneumonias in Wuhan, China.^1^ From that on, several diagnostic procedures came onto the market including highly sensitive quantitative RT-PCR methods for detecting SARS-CoV-2 out of oral and nasopharyngeal swabs as well as sputum or bronchoalveolar lavage (BAL).^2^ Despite immediate identification and characterization of the new coronavirus SARS-CoV-2,^1,3^ none of the current concepts and diagnostic algorithms were able to get the current pandemic under control. SARS-CoV-2 acts like a sniper behind the lines. Approximately 80% of infected individuals are completely asymptomatic^4^ including cases with high viral load, also designated as "Superspreaders”.^5^ But also in COVID-19 cases with clinical manifestations, SARS-CoV-2 can be transmitted two days before first symptoms occur.^6^ These circumstances clearly demonstrate why early and reliable diagnosis of COVID-19 remains a major challenge.

There is growing evidence for a transmission of SARS-CoV-2 via aerosols.^7-9^ However, only a few studies have been published to date. Liu et al. reported SARS-CoV-2 RNA concentrations in different areas of two hospitals in Wuhan.^10^ Samples were collected on styrene filter cassettes followed by a two-step RT-PCR protocol using digital droplet PCR (ddPCR). Other studies demonstrated viable SARS-CoV-2 in single hospital rooms of infected patients^11,12^ and in isolation units^13^ collected by bioaerosol samplers. Moreover, Nissen et al.^14^ were able to detect SARS-CoV-2 RNA in central ventilation systems distant from patient indicating that airborne SARS-CoV-2 can be transported over long distances. Therefore, long-range airborne transmission should be taken into consideration as a possible route of infection, even in emerging SARS-CoV-2 variants with increased infectivity.

The current outbreak of the SARS-CoV-2 B.1.1.7 lineage in the UK^15^ underlines the necessity of efficient strategies for airborne surveillance. In addition, early detection of emerging variants will be of major importance as first SARS-CoV-2 variants with an escape from neutralizing antibodies have been recently detected^16^. Positive selections among immunized individuals might drive the evolution of SARS-CoV-2 toward lineages with a partial or full resistence to current vaccination strategies.

Detection of viable SARS-Cov-2 in aerosols requires both, an effective method for sample collection, and a high-sensitive amplification procedure. Herein, we describe a simple and reliable method for the quantification of SARS-CoV-2 RNA in aerosols. Such a COVID-19 airborne surveillance could have a wide range of applications including early detection and surveillance of emerging SARS-CoV-2 variants.

## Methods

### Study design

The effects of room ventilation on SARS-CoV-2 RNA concentrations in aerosols were investigated under different conditions. Initially, measurements were performed in several rooms in two COVID-19 isolation units. The risks of viral contamination as well as the intensity of ventilation were estimated for each room. Subsequently, three locations of general interest were investigated in a high endemic area (State of Baden-Württemberg, Germany): a concert hall, an outpatient endoscopy facility, and a shopping mall.

### Procedures

Aerosol sample collections were performed by using simple, non-powered cold traps (Aeroprotektor Twin Tower, Scientifixx, Germany). The cold trap consists of two standardized 350 mL cold packs covered by a removable stainless-steel surface. The cold packs are frozen overnight in a standard freezer (−20°C). In order to prevent any contamination, the covered cold packs are frozen and transported in sealed plastic bags. At the point of interest, the frozen and covered cold packs are vertically fixed in a metal box collecting all condensed water (1-10 mL within 4-6 hours). 200 µl of condensed water is then pipetted into a nuclease free microcentrifuge tube for further analysis. Alternatively, the collecting box can be sealed containing the thawed cold packs, fixing elements, and the condensed water for immediate transportation to a specialized laboratory.

### Laboratory methods

Prior to nucleic acid amplification, SARS-CoV-2 RNA was isolated from 50 µl condensed water (Viral Xpress Kit, Merck Millipore, Darmstadt, Germany) and diluted in 50 µl AE Elution Buffer (5 mM TRIS/HCl pH 8.5, Macherey-Nagel, Düren, Germany). Alternatively, condensed water was briefly centrifuged for 1 minute at 10.000 g and pipetted directly into the qRT-PCR reaction mix. For SARS-CoV-2 quantification, 2 µl template (isolated RNA or centrifuged condensed water) was added to a 8 µl of ready-to-go CTT one-step qRT-PCR mix containing SARS-CoV-2 specific primers (targeting the RdRP gene) according to WHO criteria (Scientifixx, Gaggenau, Germany). The SYBR green based quantitative RT-PCR was performed according to manufacturer’s instructions for 40 cycles within 45 minutes including melting curve analysis on LightCycler™ 1.5/2.0 systems (Roche Diagnostics, Mannheim, Germany). A 5-log standard curve was generated by 10-fold dilutions of SARS-CoV-2 RNA isolated as described above from ATCC-VR-1986HK heat inactivated 2019-nCoV/USA-WA/2020 strain (LGC Standards, Wesel, Germany), and a 3-log dilution was used as positive control. SARS-CoV-2 Standard (Exact Diagnostics, Fort Worth, TX, USA obtained by Bio-Rad Laboratories, Feldkirchen, Germany) was used as external standard in each run.

## Results

### SARS-CoV-2 airborne contamination in isolation units

Aerosol sample collection was performed for 6 hours within an isolation room and in the corridor next to the door of the isolation room. The isolation room was ventilated permanently by two windows on tilt. The corridor next to the isolation room was ventilated sporadically while opening the house door. However, the door between the isolation room and the corridor was open during the whole aerosol collection period of 6 hours. Quantitative RT-PCR of the aerosol collected in the isolation room was negative for SARS-CoV-2 RNA. By contrast, significant amounts of SARS-CoV-2 RNA were measured in the condensed water collected by the cold trap placed in the non-ventilated corridor next to the isolation room (Ct 32).

Comprehensive aerosol measurements were then performed in several rooms of another isolation unit. As shown in table 1, SARS-CoV-2 RNA levels in collected condensed water reached concentrations up to 10^5^/mL in non-ventilated rooms of the isolation unit. However, continuous room ventilation with 1 or 2 windows on tilt reduced airborne contamination of SARS-CoV-2 RNA in collected aerosols to concentrations of 10^4^/mL or less.

**TABLE 1.** SARS-CoV-2 RNA concentrations in aerosols collected by Aeroprotektor^1^ Twin Tower within 6 hours in an COVID-19 isolation unit.

| Day | Location | Contamination<sup>2</sup> | Room Ventilation<sup>3</sup> | SARS-CoV-2 RNA<br>(copies/mL) |
| --- | --- | --- | --- | --- |
| 1 | Room 1 (evening) | ++ | ++ | 12.000 |
| 2 | Room 1 (afternoon) | + | ++ | < 2.000 |
| 2 | Room 2 (afternoon) | - | - | **100.000** |
| 2 | Room 3 (night) | - | - | **60.000** |
| 2 | Room 4 (evening) | ++ | + | 13.000 |
| 3 | Room 2 (afternoon) | - | + | 10.000 |
| 3 | Room 4 (afternoon) | ++ | + | 29.000 |
| 3 | Room 1 (afternoon) | + | ++ | 9.000 |
| 1-3 | Negative probes |  |  | - |

<sup>1</sup> Aeroprotektor is a registered trademark of Scientifixx Corp., Gaggenau, Germany
<sup>2</sup> ++ = contamination > 2 hours, + = contamination 1-2 hours, - = contamination < 1 hour
<sup>3</sup> ++ = permanent ventilation with 1-2 windows on tilt, + = intermittent ventilation, - = no ventilation

### SARS-CoV-2 spreading in a concert hall

Within a 4.000 sqft concert hall 10 cold traps were placed around an orchestra and on audience rows 3 and 5 during a 3 hours rehearsal. In one of these 10 cold traps we found measurable amounts of SARS-CoV-2 RNA (6.000 copies/mL). The contaminated cold trap was located on a loudspeaker next to the radiator at the outer wall of the concert hall.

### SARS-CoV-2 airborne contamination in an outpatient endoscopy facility

Between October 2020 and January 2021, aerosols were randomly collected by cold traps over a period of 6 hours in 2 endoscopy rooms, at the patient reception, and in a doctor’s room. In the first phase of the experiment, 12 measurements were performed. Significant amounts of SARS-CoV-2 RNA were detected in 6 collected aerosol samples. The highest SARS-CoV-2 concentration (12.000 copies/mL) was found in an endoscopy operation room, although the central ventilation system exclusively supplies outdoor fresh air avoiding any recirculation. In the second phase of the experiment, additional fresh air ventilation was initiated. As often as possible a window was on tilt in each room. Despite the additional fresh air ventilation, 4 out of 7 follow-up aerosol collections remained positive for SARS-CoV-2, but RNA concentrations were on a low level (< 2.000 copies/mL). Finally, additional fresh outdoor air ventilation was further intensified by completely opening the window between each patient in both endoscopy operation rooms. Follow-up measurements did not reveal any detectable SARS-CoV-2 RNA in the collected aerosols.

### Identification and elimination of COVID-19 hotspots in shopping malls

On a Friday 14 cold traps were placed in a shopping mall from 10 am to 3 pm. Points of interest were highly frequented areas (escalators, kiosk, checkout, and fitting room at the ground level; 3 checkouts on the 2^nd^ floor; refrigerated section, checkout as well as meat and fish counter on the 3^rd^ floor), a non-ventilated store at the ground level, and 2 staff areas (break room and administrative department). Out of these 14 cold traps, 5 contained significant levels of SARS-CoV-2 RNA. Surprisingly, the highest SARS-CoV-2 concentration (5.400 copies/mL) was found on the ground level between the escalators. The cold trap at the kiosk located next to the escalators was also positive for SARS-CoV-2 (2.000 copies/mL). Moreover, SARS-CoV-2 RNA was found in 1 out of 3 checkout areas on the 2^nd^ floor as well as on the 3^rd^ floor next to the fish counter. Both cold traps contained 2.000 copies/mL SARS-CoV-2 RNA and were located near to an inflow of the central ventilation system (2-3 meters, 7-10 ft). Another potential indoor hotspot was identified within the non-ventilated area on the ground floor. Collected aerosols revealed comparable amounts of SARS-CoV-2 RNA (2.000 copies/mL).

Follow-up measurements next Monday were performed in order to exclude any contamination of the central ventilation system. Cold traps were placed directly under 2 inflows and 2 outflows on the 3^rd^ floor. None of these cold traps contained significant amounts of SARS-CoV-2 contaminated aerosols. Control measurements at the fish counter and within the non-ventilated area at the ground level were also negative for SARS-CoV-2 RNA. However, the cold trap between the escalators at the ground level remained positive. Most likely due to the lower number of customers on a Monday, we found only half of SARS-CoV-2 RNA in collected aerosols compared to Friday before (2.700 copies/mL).

Finally, a fashion store located on 4 floors was tested for SARS-CoV-2 RNA aerosol contamination on a Friday. Compared to the shopping mall, customer frequency within the fashion store is generally less than 25%. Cold traps were placed at areas with a high customer frequency (checkout areas, escalator, fitting rooms) or less ventilated zones. However, none of the nine cold traps contained any significant amounts of SARS-CoV-2 RNA.

## Discussion

The role of SARS-CoV-2 airborne spreading in the current pandemic is still under debate, although there is growing evidence that susceptible individuals might be infected by this route of transmission^7-9^. During first SARS outbreak in 2003, Yu et al. analyzed routes of transmission among 187 cases in the Amoy Gardens housing complex.^17^ The authors concluded a three-dimensional spread of virus-loaded aerosols. Contaminated droplets and aerosols entered the air shaft carrying them upward into other apartment units. Such a route of airborne spreading is mediated by the buoyancy (chimney) effect. Recently, Kang et al. observed a similar effect for SARS-CoV-2 in a high-rise building.^18^ Most likely through drainage pipes in their master bathrooms, 3 families living in vertically aligned flats were infected by an identical SARS-CoV-2 strain.^19^ Our measurements within the concert hall also suggest a buoyancy effect on spreading kinetics of SARS-CoV-2. One single cold trap contained SARS-CoV-2 RNA at a significant concentration. However, this cold trap was located above the radiator at the exterior wall of the concert hall. The ascending flow of the heated air at the exterior wall generates a typical air circulation around the room and leads to a buoyancy effect above the radiator. Therefore, contaminated aerosols may circulate from the bottom of the hall towards the radiator at the exterior wall. These air circulation und buoyancy effects might also explain why contaminated aerosols did not reach the remaining 9 cold traps within the concert hall.

Interestingly, measurements within the shopping mall revealed similar results. Highest concentrations of SARS-CoV-2 RNA in collected aerosols were found between the escalators at the ground level, a zone predisposed for the buoyancy effect. Despite this location, contaminated cold traps were only found next to the inflow of the central ventilation system and in a non-ventilated area. Such non-ventilated areas are generally regarded as high-risk zones for the transmission of COVID-19. Our measurements within isolation units strongly support such a hypothesis. In addition, our findings within an endoscopy outpatient facility clearly demonstrate the necessity of intense fresh outdoor air ventilation in the current pandemic, even in buildings supplied by central ventilation systems avoiding any recirculation of SARS-CoV-2 contaminated air.

The identification and characterization of SARS-CoV-2 spreading kinetics and indoor hotspots are of major importance avoiding potential airborne transmission by emerging variants with an increased infectivity. Assuming the correct position of the device, SARS-CoV-2 biomonitoring by aerosol collection might enable the identification of infected groups of people as well as an early detection and surveillance of emerging SARS-CoV-2 variants even at public places. However, collecting viable SARS-CoV-2 aerosols is a rather sophisticated process requiring saturation of small virus particles prior to condensation. The BioSpot-VIVAS™ Bioaerosol Sampler has been recently demonstrated as an effective tool collecting viable SARS-CoV-2 by encapsulating airborne particles into liquid droplets followed by a deposition onto a liquid surface^11^. The method presented herein is comparably simple but the underlying mechanisms of SARS-CoV-2 collection is yet unknown. However, saturation of small virus particles into droplets capable for condensation most probably also occur. Within the first hour of the collection period, the cold traps do not contain any SARS-CoV-2 particles (data not shown). Subsequent SARS-CoV-2 aerosol saturation and condensation obviously need 2-3 hours mediated by ambient temperatures and wet surfaces which are maintained by thawing of the cold packs.

Although this method needs further optimization, it can be readily used for SARS-CoV-2 airborne surveillance. The procedure is easy to handle, cost efficient and suitable to be used worldwide even in emerging and developing countries.

## Data Availability

All data referred to in this manuscript are available on request from the corresponding author.

## Notes

### Competing Interest Statement

The authors have declared no competing interest.

### Funding Statement

There is no funding support in the work presented. The authors and their institution did not receive any payment or services from a third party for any aspect of the submitted work.

### Author Declarations

As discussed prior to submission, clinical studies on this topic are in progress and are currently evaluated by the federal ethics committee of the physicians board of the State of Baden-Wuerttemberg, Germany (F-2020-176). Due to the potential impact of the present data on human health we submit this paper prior to a a final decision about the design of our clinical studies. The present paper describes SARS-CoV-2 airborne surveillance using a simple aerosol collection procedure and does not contain any clinical data or informations about health conditions.

